# Buruli ulcer Community Health Education and Medical Screening (BU-CHEMS) in Ga South District, Ghana

**DOI:** 10.1101/2024.07.31.24311162

**Authors:** Charles A. Narh, Edwin Tetteh, NSPA-NMIMR

## Abstract

**Background:** The National Service Personnel Association of the Noguchi Memorial Institute for Medical Research (NSPA-NMIMR) carries out an annual community outreach project comprising health education and medical screening for some of the diseases that the institute works on. Therefore, the NSPA 2009/2010 group (40 personnel) conducted Buruli ulcer community health education and medical screening (BU-CHEMS) in the Ga South District of Ghana in July 2010.

**Method:** BU is caused by *mycobacterium ulcerans*, and starts as a painless nodule, which can progress to ulcer, particularly on the upper and lower extremities. BU is often associated with witchcraft in some Ghanaian communities, and as a result, some patients reluctantly seek medical treatment. Therefore, prior to the screening program, the NSPA and medical staff from the Obom Health Centre showed video documentaries on BU as a way of educating and dispelling myths about the disease. This was followed by screening for nodules and ulcers among 2,500 people, mostly children in primary schools, residing in four endemic communities in the Ga South District. Other medical screening activities included blood group/pressure tests, Body Mass Index (BMI) and body temperature checks.

**Results and conclusion:** Suspected cases of BU (N=33) ranging from nodules, plagues, oedema and ulcers of the disease on various parts of the body including the lower and upper extremities were identified, and samples were sent to NMIMR for PCR confirmation. All the PCR-positive cases (78%), including children (<15 years, 88%) were referred to the Obom Health Center for clinical treatment. The BU-CHEMS organized by NSPA 2009/2010 group (with sponsorship from corporate organizations) contributed to NMIMR mandate: improving the health and wellbeing of Ghanaians and mankind through focused and relevant quality biomedical research, human resource development and support of national public health activities.

## INTRODUCTION

Buruli ulcer (BU), a disease caused by *Mycobacterium ulcerans* (MU) is one of the most neglected but treatable tropical diseases. MU belongs to the same family of mycobacteria that causes tuberculosis, but BU has received less attention. BU is endemic in more than 30 countries, occurring mostly West Africa and Southeastern Australia [1]. In some endemic communities in Ghana, BU is associated with witchcraft and is known by local names such as ‘‘Odontihela’’ (describing the cotton wool appearance associated with the fatty necrosis), ‘‘Aboa gbonyo’’ (dreadful disease), and ‘‘Ashanti Asane’’ (meaning the disease originated from the Ashanti region) [2]. Infection leads to extensive destruction of skin and soft tissue with the formation of large ulcers usually on the legs or arms.

In Ghana, public health focus has largely been on early detection and complete excision of nodule with antibiotic treatment, and surgery with skin grafts for ulcers to prevent permanent disability [3]. In Ghana a few hospitals such as the Amasaman District Hospital and the St. Martins Catholic Hospital, and some Community Health Centres including the Obom Health Centre in the Ga South District of the Greater Accra region offer free medical care to BU patients. Most of the BU endemic communities are rural and lack good drinking water. In addition, they are superstitious about the disease, with myths associating BU with witchcraft.

Therefore, as part of the annual community engagement activities of the Noguchi Memorial Institute for Medical Research, the National Service Personnel Association (NSPA) 2009/2010 group of 40 Research Assistants conducted voluntary Buruli ulcer community health education and medical screening (BU-CHEMS) to educate and raise awareness about BU and to screen for cases in four endemic communities in the Ga South District of Ghana. This report presents a summary of the CHEMS activities conducted by the group (see full list of members in Appendix, Table 1).

**Table 1:**
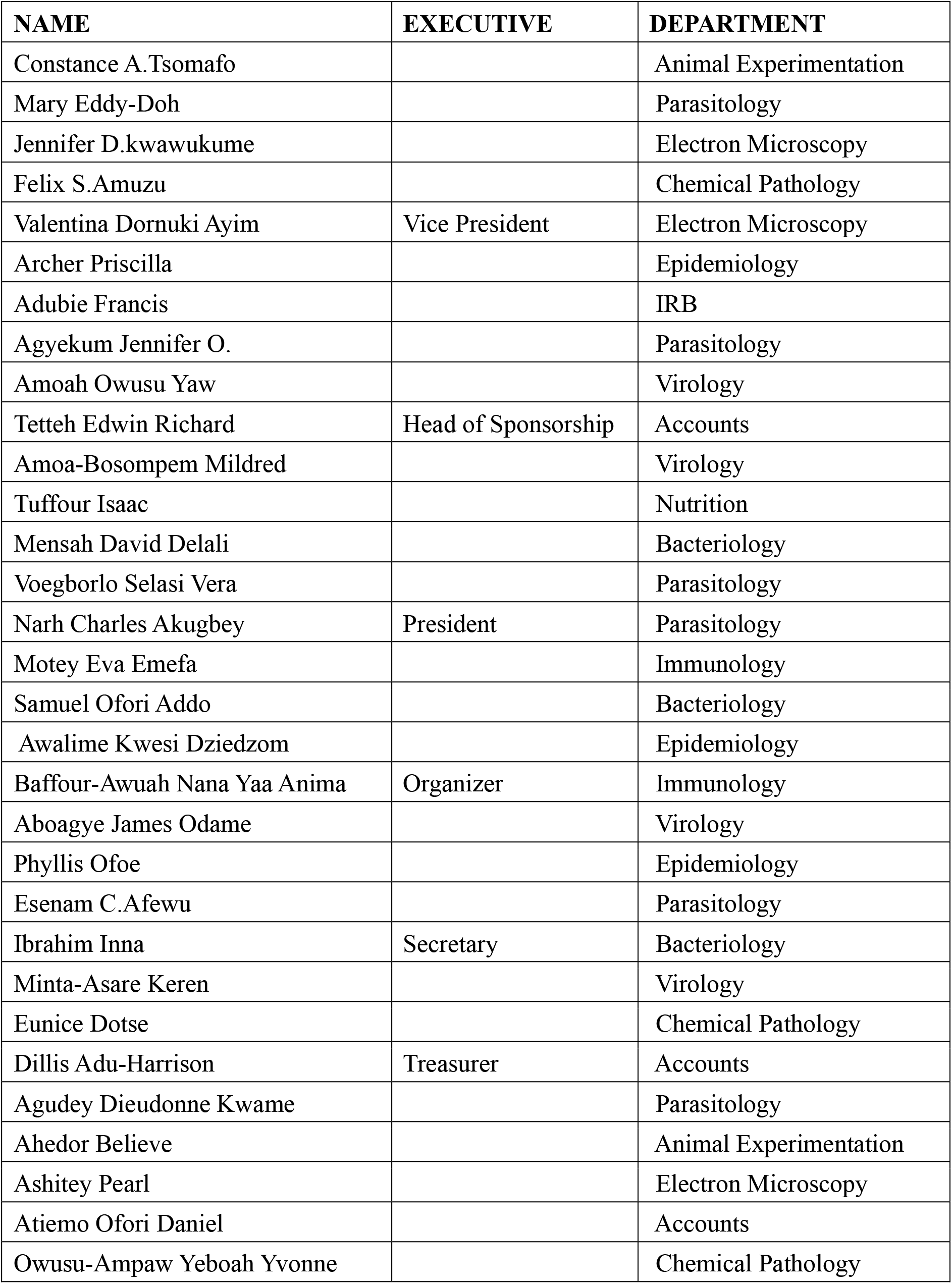
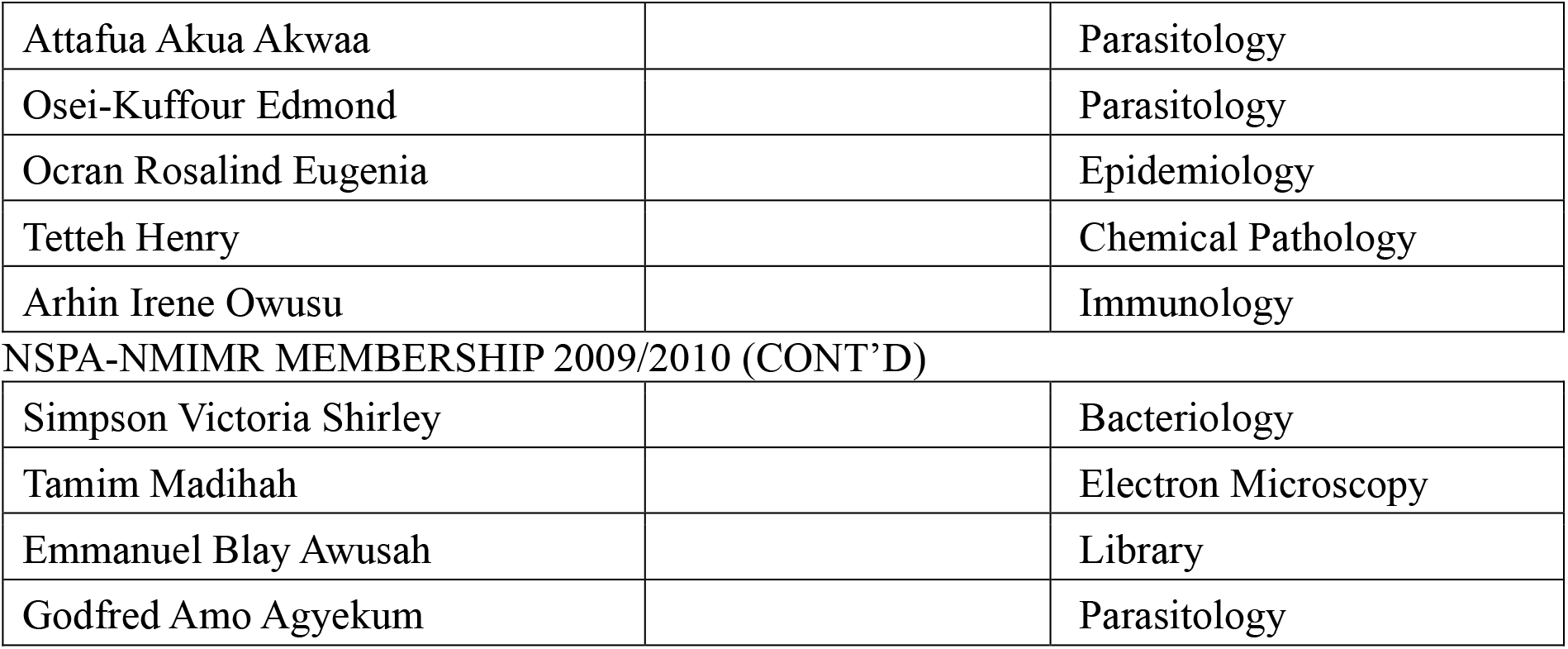
NSPA-NMIMR MEMBERSHIP 2009/2010.

| NAME | EXECUTIVE | DEPARTMENT |
| --- | --- | --- |
| Constance A.Tsomafo |  | Animal Experimentation |
| Mary Eddy-Doh |  | Parasitology |
| Jennifer D.kwawukume |  | Electron Microscopy |
| Felix S.Amuzu |  | Chemical Pathology |
| Valentina Dornuki Ayim | Vice President | Electron Microscopy |
| Archer Priscilla |  | Epidemiology |
| Adubie Francis |  | IRB |
| Agyekum Jennifer O. |  | Parasitology |
| Amoah Owusu Yaw |  | Virology |
| Tetteh Edwin Richard | Head of Sponsorship | Accounts |
| Amoa-Bosompem Mildred |  | Virology |
| Tuffour Isaac |  | Nutrition |
| Mensah David Delali |  | Bacteriology |
| Voegborlo Selasi Vera |  | Parasitology |
| Narh Charles Akugbey | President | Parasitology |
| Motey Eva Emefa |  | Immunology |
| Samuel Ofori Addo |  | Bacteriology |
| Awalime Kwesi Dziedzom |  | Epidemiology |
| Baffour-Awuah Nana Yaa Anima | Organizer | Immunology |
| Aboagye James Odame |  | Virology |
| Phyllis Ofoe |  | Epidemiology |
| Esenam C.Afewu |  | Parasitology |
| Ibrahim Inna | Secretary | Bacteriology |
| Minta-Asare Keren |  | Virology |
| Eunice Dotse |  | Chemical Pathology |
| Dillis Adu-Harrison | Treasurer | Accounts |
| Agudey Dieudonne Kwame |  | Parasitology |
| Ahedor Believe |  | Animal Experimentation |
| Ashitey Pearl |  | Electron Microscopy |
| Atiemo Ofori Daniel |  | Accounts |
| Owusu-Ampaw Yeboah Yvonne |  | Chemical Pathology |
| Attafua Akua Akwaa |  | Parasitology |
| Osei-Kuffour Edmond |  | Parasitology |
| Ocran Rosalind Eugenia |  | Epidemiology |
| Tetteh Henry |  | Chemical Pathology |
| Arhin Irene Owusu |  | Immunology |
| Simpson Victoria Shirley |  | Bacteriology |
| Tamim Madihah |  | Electron Microscopy |
| Emmanuel Blay Awusah |  | Library |
| Godfred Amo Agyekum |  | Parasitology |

## METHODS

### BU media outreach

Prior to the commencement of the project, NSPA organized a conference dubbed ‘‘ Buruli ulcer conference’’ on 18 June 2010 at the NMIMR Conference Hall. The main aim of this was to educate staff and general public about BU and acquaint staff with the project. The occasion was also used to appeal for funds to carry out the project. In attendance was members of the University of Ghana community, and general public including the media (telecasted on TV3, GTV, NET 2, Radio Univers and GBC Radio). See Appendix, Table 2 for list of sponsors.

**Table 2:**
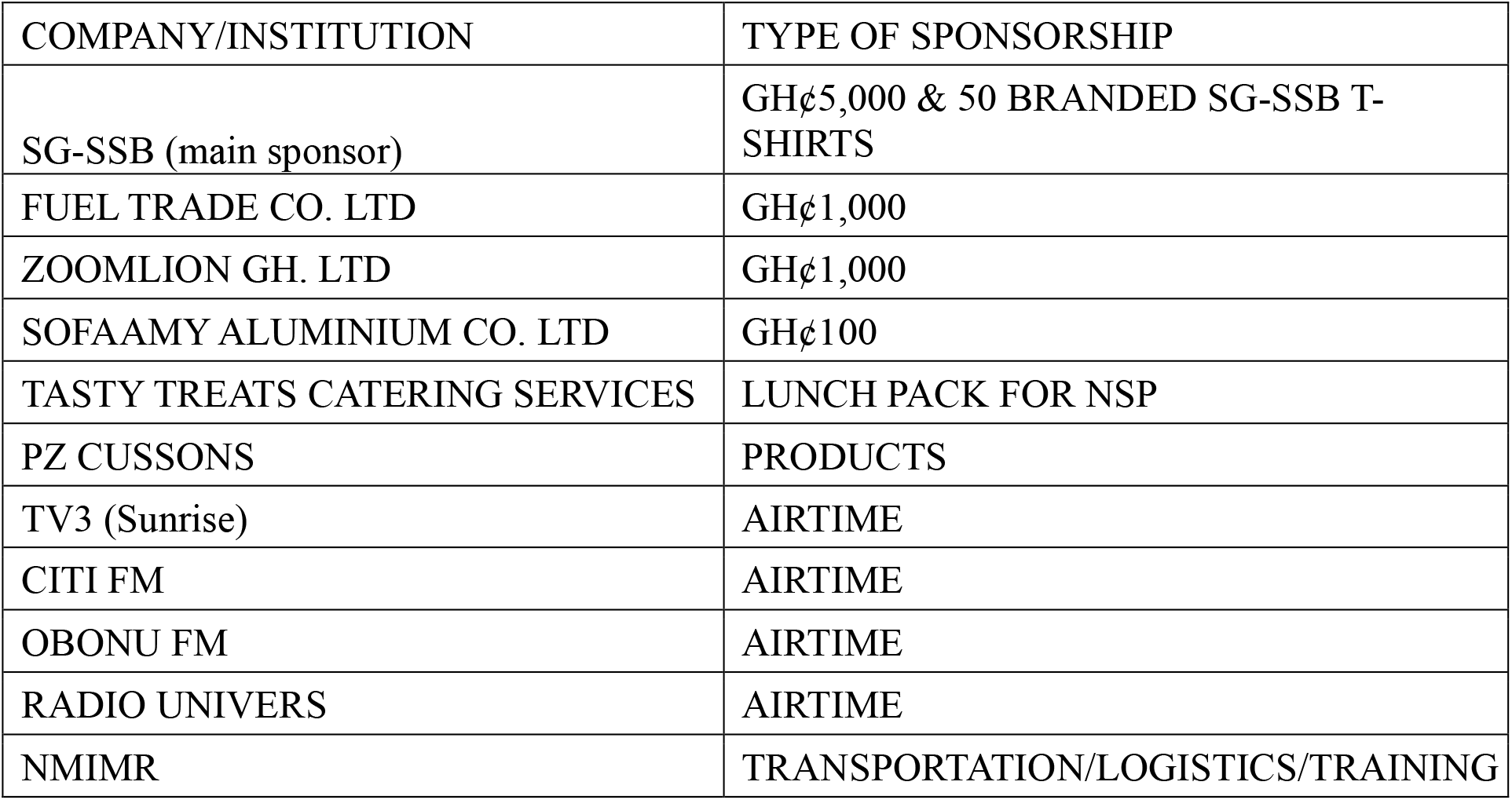
SPONSORSHIPS.

The conference was chaired by Prof. George Armah (NMIMR), with speakers including Dr. Edwin Ampadu (Director of the National Buruli ulcer Control Programme) who talked about the pathology and epidemiology of the disease as well as the current strategies by the nation to control and eradicate the disease. And Dr. Lydia Mosi (Research Fellow/Lecturer, West African Centre for Cell Biology of Infectious Pathogens/WACBIP, University of Ghana) who spoke about ongoing international research efforts to find new therapeutics including vaccines for the disease.

Additionally, the NSPA team led by Charles Narh (President of the Association) conducted media education about BU and the project and led sponsorship campaigns to raise funds to support the project. These media engagement activities were also telecasted on national TV as mentioned above.

### Field work and BU-CHEMS

Between April-June, we undertook community entry to seek permission from the community leaders (Chiefs and District Health Directorates) to conduct the project in four communities – Domiabra, Ashalaja. Balagonno and Danchira in the Ga South District. The project was also approved and supported by the National Buruli ulcer Control Programme and NMIMR. The project was carried out in two phases.

Phase A comprised of general health and BU education and awareness, conducted 21-22 July 2010. This was done by showing WHO-certified BU documentaries to residents in the four communities. The documentaries (“video show”) were shown on large screens during the early evenings at the community market squares or playgrounds (>300 attendees per show per community). They were produced in English and therefore had to be interpreted in the local dialect – Ga language for easy understanding. The NSPA team engaged the services of eight community health workers and disease control officers to help mobilize the community for the BU documentaries and screening activities.

Phase B (Figure 1) comprised of general health checks and BU screening, conducted 22-23 July 2010. Five stances were mounted for BU screening, BMI, blood pressure, blood group and body temperature. A total of 2,500 participants including 2,200 primary school children (<15 years) and 300 adults (≥ 18 years) were screened for BU based on physical examinations that looked for nodules, oedema, papules or ulcers. Fine needle aspirates of nodules/oedema and swabs of ulcers were taken from suspected BU cases for PCR confirmation NMIMR as extensively described elsewhere [4].

**Figure 1:**
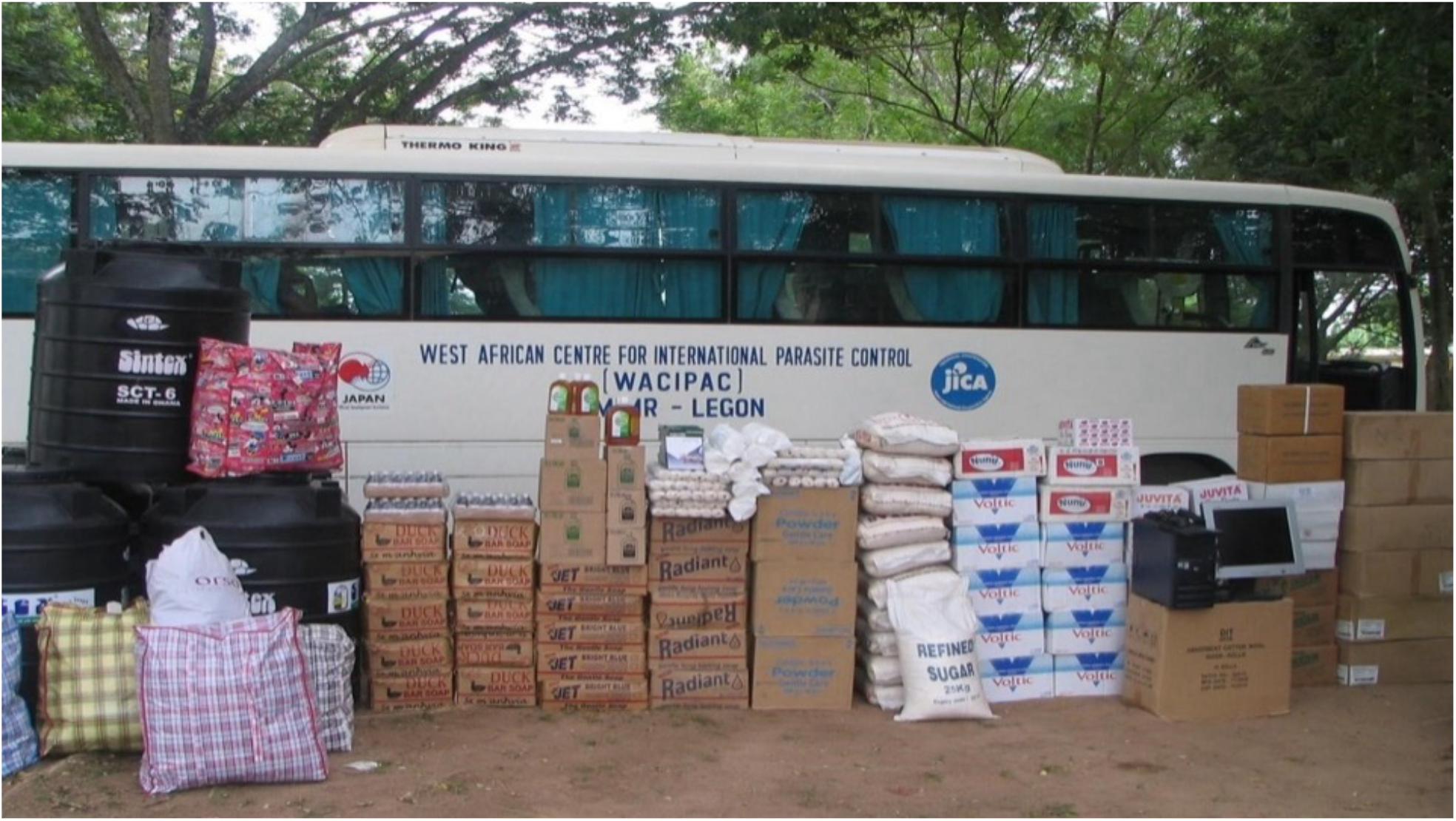
Buruli ulcer Community health education and medical screening (BU-CHEMS). Medical items donated to the Obom Health Centre for the management of Buruli ulcer cases. The medical supplies included gauze, Vaseline® gauze, gloves, Dettol®, soaps, cotton wool and methylated spirit. Other donations included bags of rice and sugar, oats, Milo®, biscuits and drinks, Voltic® bottled and sachet water, used clothes and a desktop computer. The NSPA also donated a Sintex polytank (to store drinking water) to each of the four schools and to the Health Center. About 2,500 community members including 2,200 children (< 15 years) in primary schools and 300 adults in the four endemic communities were screened for BU. Confirmed positive cases were treated at the Obom Health Centre.

#### Participant data collection

The NSPA team also administered questionnaires to collect data on participants’ demographics and knowledge about BU. The following questions were asked; Have you ever heard about Buruli ulcer? Where did you hear it from? Do you know what causes Buruli ulcer? Will you go to the health center if you had Buruli ulcer? If a friend had Buruli ulcer, will you play or study with that person?

#### Project approval

The project was supported by Noguchi Memorial Institute for Medical (NMIMR) University of Ghana), the National Buruli Ulcer Control Programme (NBUCP, Korle-Bu Hospital) and the Obom Health Center. These institutions provided written approval for the conduct of the project. The sample collection was embedded into a large BU diagnosis study, with ethics approval from the NMIMR Institutional Review Board [5] [6]. All participants consented prior to participating and having their samples taken for BU test confirmation at NMIMR. Refreshment was provided for all participants.

## RESULTS

### Demographics

Due to logistical challenges, the questionnaires were randomly administered to 50 participants. Of this number, 85% had heard about the disease (mainly from home or school). Over 50% of respondents had no idea of the cause of the disease. Among those who knew the cause, 85% attributed it to microorganisms while 15% attributed it to either witchcraft or punishment from a deity. Also, over 90% of respondents said they would seek medical treatment. Regarding the question of social contact, 58% would not play with affected victims for fear of infection.

### Laboratory confirmation of suspected Buruli ulcer cases

2,200 students from four schools participated ant about 300 adults from the respective communities were also screened. Only those with clinical manifestations of the disease had their blood and swab samples taken for laboratory confirmation. Laboratory test, Polymerase Chain Reaction (PCR) was performed at the Bacteriology Department, NMIMR (with due acknowledgement to Prof. Dorothy Yeboah-Manu group). In all, 33 samples were analyzed – 78.8% and 21.2% were positive and negative, respectively, for the MU. Most of the victims had single lesions with only a few having multiple (≥2) Category III lesions. All the cases seen were ulcerative. There were no recorded cases from Balagonno but 88% of these positive cases were children below 15 years of age (all children were primary school students) with the remaining 12% being 18-34 years inclusive. The positive cases comprised of females (42.3%) and males (57.6%).

For BMI the cutoffs were < 19 kg/ m^2^ underweight, 19-25 kg/ m^2^ normal, 25.1-28 kg/ m^2^ mild obesity, 28.1-32 kg/ m^2^ moderate obesity, 32.1-35 kg/ m^2^ severe obesity and > 35.1 kg/ m^2^ morbid obesity (based on the Standard Treatment and Guidelines, Ministry of Health, Ghana, 2000). Of the 52 screened, the majority had normal BMI (56.9%), Notably, 1.7%, 17.2%, 5.2%, 12.1% and 6.9% were underweight, mild, moderate, severe and morbid for obesity. Participants had their readings explained to them and given appropriate medical counselling to see their doctor.

For Blood pressure, out of a total of 72 people (mostly elderly, 40 years +), 4.2%, 70.3%, 13.9% and 11.1% had low, ideal, pre-high and high BP respectively.

For blood group tests: Of the 424 participants screened, 194 (45.9%), 13 (3.1%), 93 (21.9%), 5 (1.2%), 79 (18.7%), 17 (4.0%), and 22 (5.2%) were O+, A-, A+, B-, B+, AB-, AB+ and O-, respectively.

## DISCUSSION

The project was carried out at Obom in the Ga South District of the Greater Accra region from the 21-23 July 2010. Four communities within Obom; Domiabra, Ashalaja, Danchira and Balagonno were the beneficiaries of the project. The project aimed at educating these communities about Buruli ulcer (BU) and screening them for the disease. These communities were reported by the National Buruli ulcer Control Programme (NBUCP), Korle-BU Teaching Hospital as being endemic for the disease. Therefore, Our activities were to complement efforts by the NBUCP and other Non-Governmental Organizations (NGOs) to control BU in these communities and Ghana as a whole. Furthermore, the association also saw the need to assist the Obom Health Center with some medical supplies to cater for BU patients.

The educational campaigns were successful as the residents were already gathered at the playgrounds each time we went in. This was due to the fact that prior to the project, the communities were sensitized about the project via the community health volunteers and the ‘‘gongon beater’’. The BU documentaries were interpreted in the local dialect (Ga). Questions about the disease were properly answered and all issues concerning the documentaries were adequately clarified in the Ga language. We observed that most (85%) of the inhabitants were a bit knowledgeable about the disease. The Ministry of Health and other NGOs had carried out BU educational programs in these communities during previous years, which might explain the latter observation. However, there were other issues, such as the causative organism and risk factors for infection, that we had to explain to them during BU documentary shows and screening activities.

The questionnaire results showed that education plays a significant role in BU control programs. Therefore, more resources should be channeled into educating children living in endemic communities. The schools and homes could be empowered to spearhead such educational campaigns as these are the two places where the children spend much of their time. There is also the need to bring primary healthcare to the doorsteps of people living in endemic communities. Although treatment is free, most BU patients found it difficult to commute between their homes and the Health Centers due to poor condition of roads, lack of transportation fare, stigmatization, etc. Furthermore, the Health Centers could be upgraded to have BU wards. This may ease transportation problems and enable physicians to properly monitor patients in admission.

Among the confirmed cases (n=26), 88% were children below 15 years having different clinical manifestations of the disease. Although, we screened mostly children, this result suggests that children leaving in endemic areas are vulnerable to infection. This could be due to several factors – children run errands to the riverside to fetch water for domestic chores. Furthermore, they also sometimes go to the riverside for a swim. These activities could be risk factors [2] which predispose these children to MU infection and could therefore explain the high percentage of cases in this cohort. However, the mode of transmission is still not clear. Studies in this cohort (children below 15 years) in endemic areas could provide some answers to the mode of transmission.

All identified cases were ulcerative, and patients had their wounds dressed by trained BU health personnel from the Obom Health Centre. This shows that most patients are either unaware of BU symptoms and therefore delay in seeking medical treatment or they purposefully refuse to seek early treatment due to the painless nature of symptoms. Others were not aware that BU treatment was free. There is therefore the need to recruit more health workers like community nurses who have been trained adequately in BU healthcare and can take BU education and medical assistance to the doorstep of at-risk community members in endemic areas. Furthermore, community Health Centers can be upgraded and equipped with adequate resources to provide a holistic BU-healthcare delivery.

The association also provided other free screening services like BMI, blood pressure (BP), blood group tests and body temperature checks. Majority of the participants, >70% had BP within the ideal/normal range. For the BMI, it was realized that 56% were within the normal range. These estimates are good health indicators, which may suggest that the participants had healthy lifestyles. Most of these people we screened and interacted with were full-time farmers who walked long distances (>15km) to their farmers. A few were engaged in enterprises which required them to commute long distances. Children were exempted from these tests. We assumed they were within normal range as cases of obesity and high BP are mostly seen in the elderly. The majority of participants were O+ (45.9%) for the blood group test. Each participant was given a form containing his/her name, age, sex and test results (BMI, BP, blood group and body temperature). They were advised to keep them safely for future reference. The significance of each test was thoroughly explained to each patient before and after each test. Those who had test readings outside the normal ranges were advised to seek medical care.

## Data Availability

All data produced in the present work are contained in the manuscript

## RECOMMENDATION

It is the association’s wish that this project be sustained by the next batch of service personnel. There are some other communities within Obom which are endemic with the disease and have had no outreach program extended to them. In this light, the executives, on behalf of the entire membership of NSPA-NMIMR 2009/2010 would be happy if NMIMR administration continues its support of the CHEMS activities.

## DECLARATION

This project dubbed “Buruli ulcer Community Health Education and Medical Screening (BU-CHEMS) in Ga South District, Ghana” was carried out by the National Service Personnel Association of Noguchi Memorial Institute for Medical (NSPA-NMIMR), 2009/2010 batch with permission and support from the National Buruli Ulcer Control Programme (Korle-Bu Hospital), Noguchi memorial Institute for Medical, University of Ghana, Legon and the Obom Health Center. This report was prepared by Charles Narh (President, NSPA-NMIMR) for NSPA-NMIMR 2009/2010 batch. Mr. Edwin Tetteh contributed to writing and review of the final draft. The authors declare no competing interest.

## SPONSORSHIP AND TRANSPARENCY STATEMENT

The association was able to donate items worth GH¢ 6,500 to the Obom Health Center. Much of the funds used for the project came from the Societe Generale Ghana (SG-SSB) bank, i.e. SG-SSB, the main sponsor. The bank sponsored the association with GH¢ 5,000 and 50 branded SG-SSB T-shirts. Fueltrade Oil Ltd and Zoomlion GH Ltd donated GH¢1,000 each. Voltic GH Ltd gave the association 20 cartons of 500ml water and 20 bags of sachet water, PZ Cussons supported with some of their products and Tasty Treat Catering services gave the association take-away lunch packs for the 40 service personnel for two days. Additionally, NMIMR staff donated used clothes. Lastly, NSPA members contributed GH¢1,925 aside monthly dues for the project. The association also acknowledges contributions in cash and/or in kind from staff of NMIMR towards the project. The media stations TV3, GTV, Obonu fm, Citi fm, Radio Univers and Net 2 TV are duly acknowledged.

It was our hope that we would be able to raise GH¢21,000 for this project. Unfortunately, we could not. Most of the companies we wrote to for sponsorship had other priorities. For some companies, the BU-CHEMS project was not within their scope of corporate social responsibilities. Approximately, about 90% of sponsorship funds was used for goods/items and services for the project. The expenses incurred by NSPA including administration services for sponsorship advocacy were paid using our monthly dues and contributions. The association had GH¢859.20 left in its accounts, which it credited to the next administration for sustenance and furtherance of this project.

## ACKNOWLEDGEMENT

The executives and the entire membership of NSPA-NMIMR, University of Ghana, thank the Almighty God for His divine guidance and providence for the success of this project. Sincere thanks and gratitude to Prof. Alexander Nyarko (Director, NMIMR) and Prof. Kwadwo Koram (Deputy director, NMIMR) for their unflinching support.

Furthermore, we thank Dr. Ben Gyan (Head, Immunology), the NSPA-NMIMR 2009/2010 patron, for his mentorship. Furthermore, we acknowledge efforts from Dr. Dorothy Yeboah Manu and Dr. Edwin Ampadu who played both supervisory/advisory roles on the BU-CHEMS project. We thank Dr. Lydia Mosi, Dr. Charles Quaye, Linda Daisie and Cherie McCown for the research and technical supports/advice. We thank NMIMR staff.

The project was supported by Noguchi Memorial Institute for Medical (NMIMR) University of Ghana), the National Buruli Ulcer Control Programme (Korle-Bu Hospital), the Obom Health Center and the Societe Generale Ghana (SG-SSB) bank, our main sponsor. The media stations TV3, GTV, Obonu fm, Citi fm, Radio Univers and Net 2 TV are duly acknowledged.

### APPENDIX

#### NSPA-NMIMR PROFILE

The National Service Personnel Association of Noguchi Memorial Institute for Medical Research (NSPA-NMIMR) is a recognized association in Noguchi for all national service personnel. It was formed by the 2007/2008 batch of service personnel. The association is governed by an executive board elected every year by the members. These includes the president, vice president, secretary, treasurer, organizer and the Head of Sponsorship who is elected by the other five executives after induction into office. The association also has a patron who sees to the general well-being of all the members and monitors matters of the association. Members pay monthly dues for the administrative running of the association. As part of our end of service celebration, the association carries out a charity project to help the less privileged in the society. Past projects have been medical screening for the inmates of the Ho Prisons (2008) and a Charity project at the Ho Leprosarium (2009). Our group 2009/2010 voted to undertake the BU-CHEMS following feasibility and endorsement by NMIMR administration.

#### CONTACT US

Noguchi Memorial Institute for Medical Research

College of Health Sciences

University of Ghana

P. O. Box LG 581

Legon-Accra

**Please note**: Charles Narh is now with Deakin University

## Notes

### Competing Interest Statement

The authors have declared no competing interest.

### Funding Statement

The project was supported by Noguchi Memorial Institute for Medical (NMIMR) University of Ghana) and the National Buruli Ulcer Control Programme (Korle-Bu Hospital) and the Obom Health Center and the Societe Generale Ghana (SG-SSB) bank, our main sponsor. The media stations TV3, GTV, Obonu fm, Citi fm, Radio Univers and Net 2 TV are duly acknowledged.

### Author Declarations

The project was supported by Noguchi Memorial Institute for Medical (NMIMR) University of Ghana), the National Buruli Ulcer Control Programme (NBUCP, Korle-Bu Hospital) and the Obom Health Center. These institutions provided written approval for the conduct of the project. The sample collection was embedded into a large BU diagnosis study, with ethics approval from the NMIMR Institutional Review Board. All participants consented prior to participating and having their samples taken for BU test confirmation at the Noguchi Memorial Institute for Medical Research, University of Ghana. Refreshment was provided for all participants.

## REFERENCES

1. Omansen, T.F., et al., Global Epidemiology of Buruli Ulcer, 2010-2017, and Analysis of 2014 WHO Programmatic Targets. Emerg Infect Dis, 2019. 25(12): p. 2183–2190.

2. Asiedu, K. and S. Etuaful, Socioeconomic implications of Buruli ulcer in Ghana: a three-year review. Am J Trop Med Hyg, 1998. 59(6): p. 1015–22.

3. Tuwor, R.D., et al., Stigma experiences, effects and coping among individuals affected by Buruli ulcer and yaws in Ghana. PLoS neglected tropical diseases, 2024. 18(4): p. e0012093.

4. Sakyi, S.A., et al., Clinical and Laboratory Diagnosis of Buruli Ulcer Disease: A Systematic Review. Canadian Journal of Infectious Diseases and Medical Microbiology, 2016. 2016(1): p. 5310718.

5. Ahorlu, C.K., et al., Enhancing Buruli ulcer control in Ghana through social interventions: a case study from the Obom sub-district. BMC Public Health, 2013. 13(1): p. 59.

6. Yeboah-Manu, D., et al., Laboratory confirmation of Buruli ulcer cases in Ghana, 2008-2016. PLOS Neglected Tropical Diseases, 2018. 12(6): p. e0006560.

